# Highly-Sensitive Lineage Discrimination of SARS-CoV-2 Variants through Allele-Specific Probe Polymerase Chain Reaction

**DOI:** 10.1101/2021.11.01.21265384

**Authors:** Jeremy Ratcliff, Farah Al-Beidh, Sagida Bibi, David Bonsall, Sue Ann Costa Clemens, Lise Estcourt, Amy Evans, Matthew Fish, Pedro M. Folegatti, Anthony C. Gordon, Cecilia Jay, Aislinn Jennings, Emma Laing, Teresa Lambe, George MacIntyre-Cockett, David Menon, Paul R. Mouncey, Dung Nguyen, Andrew J. Pollard, Maheshi N. Ramasamy, David J. Roberts, Kathryn M. Rowan, Jennifer Rynne, Manu Shankar-Hari, Sarah Williams, Heli Harvala, Tanya Golubchik, Peter Simmonds, the AMPHEUS Project, REMAP-CAP Immunoglobulin Domain UK Investigators, and Oxford COVID-19 Vaccine Trial Group

**Author notes:** To whom correspondence should be addressed: Jeremy Ratcliff, Peter Medawar Building for Pathogen Research, S Parks Rd, University of Oxford, Oxford, United Kingdom. Wellcome Centre for Human Genetics, University of Oxford, Oxford, United Kingdom.

## Abstract

**Introduction:** Tools to detect SARS-Coronavirus-2 variants of concern and track the ongoing evolution of the virus are necessary to support public health efforts and the design and evaluation of novel COVID-19 therapeutics and vaccines. Although next-generation sequencing (NGS) has been adopted as the gold standard method for discriminating SARS-CoV-2 lineages, alternative methods may be required when processing samples with low viral loads or low RNA quality.

**Methods:** An allele-specific probe polymerase chain reaction (ASP-PCR) targeting lineage-specific single nucleotide polymorphisms (SNPs) was developed and used to screen 1,082 samples from two clinical trials in the United Kingdom and Brazil. Probit regression models were developed to compare ASP-PCR performance against 1,771 NGS results for the same cohorts.

**Results:** Individual SNPs were shown to readily identify specific variants of concern. ASP-PCR was shown to discriminate SARS-CoV-2 lineages with a higher likelihood than NGS over a wide range of viral loads. Comparative advantage for ASP-PCR over NGS was most pronounced in samples with Ct values between 26-30 and in samples that showed evidence of degradation. Results for samples screened by ASP-PCR and NGS showed 99% concordant results.

**Discussion:** ASP-PCR is well-suited to augment but not replace NGS. The method can differentiate SARS-COV-2 lineages with high accuracy and would be best deployed to screen samples with lower viral loads or that may suffer from degradation. Future work should investigate further destabilization from primer:target base mismatch through altered oligonucleotide chemistry or chemical additives.

## INTRODUCTION

The ongoing evolution of SARS-CoV-2 leading to emergence of variants of concern (VoC) and variants of interest (VoI) has highlighted the need for broadly accessible methods for detecting and tracking mutations in the SARS-CoV-2 genome. Next-generation sequencing (NGS) with a collection of different methods and protocols has been largely adopted as the gold standard approach for detecting SARS-CoV-2 variants. Several targeted and untargeted whole-genome NGS methods have been deployed for sequencing SARS-CoV-2, including the tiling amplicon-based ARTIC protocol and targeted enrichment RNA-seq-based veSeq protocol.^1,2^

Tools to identify the emergence of novel SARS-CoV-2 variants and track their spatial spread are necessary to support public health interventions. Single-nucleotide polymorphisms (SNPs) such as S:D614G may be associated with increased transmissibility.^3^ VoCs Alpha (Pangolin: B.1.1.7 and Q.*), Beta (Pangolin: B.1.351), Gamma (Pangolin: P.1 and P.1.*), and Delta (Pangolin: B.1.617.2 and AY.*) have constellations of mutations that, to varying degrees, appear to impact transmissibility, severity, and evasion of vaccine-derived immune responses.^4^ Rapid identification of novel lineages through NGS is necessary for accurate characterization and risk assessment. However, recovering whole-genome sequences for samples with low viral load is challenging and RNA quality must be high, conditions that may not be met in samples taken from patients in the later phases of infection where viral loads may be low or in settings where optimal sample handling and storage is not readily available.^5,6^ In particular, high levels of RNA degradation during storage or transport of primary material or RNA extracts can severely impact the effectiveness of amplicon-based protocols such as ARTIC, the most popular SARS-CoV-2 sequencing technique,^1^ because of its dependence on the generation of 400bp amplicons in its pre-amplification step.

Herein, we describe the development and application of a polymerase chain reaction (PCR)-based, high-throughput method for SARS-CoV-2 lineage designation – allele-specific probe (ASP)-PCR - and describe its application in two patient cohorts. ASP-PCR leverages differential binding affinities of two fluorescently-labelled probes differing in the base overlapping a SNP site to designate SNPs at lineage informative locations in the SARS-CoV-2 genome. The method uses similar amplicon length (120-200 bp) and technology as real-time quantitative PCR (RT-qPCR), it is therefore more robust to degraded RNA than ARTIC and accessible to most microbiology laboratories. Here, ASP-PCR was applied to detect three SNPs in the SARS-CoV-2 genome, each highly specific and selective for VoCs and VoIs relevant to the respective trials.

The Randomized, Embedded, Multifactorial, Adaptive Platform Trial for Community-Acquired Pneumonia (REMAP-CAP) Immunological Domain trial conducted internationally but with principal patient representation from within the United Kingdom recruited 2,097 patients to a convalescent plasma trial from March 9, 2020 to January 18, 2021.^7,8^ Crucially, reports indicate the emergence of the antigenically distinct Alpha (pangolin: B.1.1.7) variant in the United Kingdom from November 2020, overlapping trial recruitment.^9^ Indeed, our previously published data indicated >50% of trial infections occurring after the second week of December were due to Alpha, reaching >80% of new infections by the end of trial recruitment.^10^ To detect and differentiate those infections caused by Alpha, the spike mutation S:D1118H was targeted by ASP-PCR. The COV003 trial is a phase III trial of the ChAdOx1 nCoV-19 vaccine in Brazil that overlapped the emergence of both the Gamma (pangolin: P.1) and P.2 variants in the country.^11,12^ For these, ASP-PCR assays were designed targeting the spike mutation S:K417T for Gamma and ORF1a:L3468V for P.2.

In both trials, we compared the ability of ASP-PCR and the veSeq NGS method to perform lineage designation. While not as widely used as the ARTIC protocol, veSeq involves bait capture rather than amplicons, and as such is more resistant to RNA fragmentation and degradation, which was necessary given the quality of RNA in the COV003 trial, and allows for robust, quantitative assessment of viral minor populations. Application of probit regression models for each technique derived from the two patient cohorts to a simulated viral load dataset clearly demonstrate the increased sensitivity of ASP-PCR over NGS for specific allele genotyping and lineage discrimination across a wide range of viral loads in both cohorts.

## RESULTS

### SNP sensitivity and specificity

To assess the suitability of different SNP targets for lineage specification in the ASP-PCR platform, global sequencing data published as part of the GISAID repository was utilized, leveraging the collation performed by Outbreak.info (https://outbreak.info).^13,14^ The sensitivity, specificity, positive predictive value (PPV), and negative predictive value (NPV) for various lineage-defining SNPs within the countries where the samples were collected was calculated (Table 1). Despite being designed when little sequencing data was available, the SNPs used in this study were shown to be robust and identify their respective lineages with very high accuracy. Within the United Kingdom, the S:D1118H SNP is present in 99.90% of Alpha infections and detection of the SNP gives a 99.93% chance that the sample represents an Alpha infection. The S:L3468V SNP proved to be overall the best SNP to target P.2 with the highest PPV of analyzed SNPs of 99.69%. For Gamma, the S:K417T SNP was not the most appropriate target and demonstrated the lowest sensitivity among the ten SNPs analyzed (93.72%), but still maintained very high PPV (99.42%).

**Table 1).**
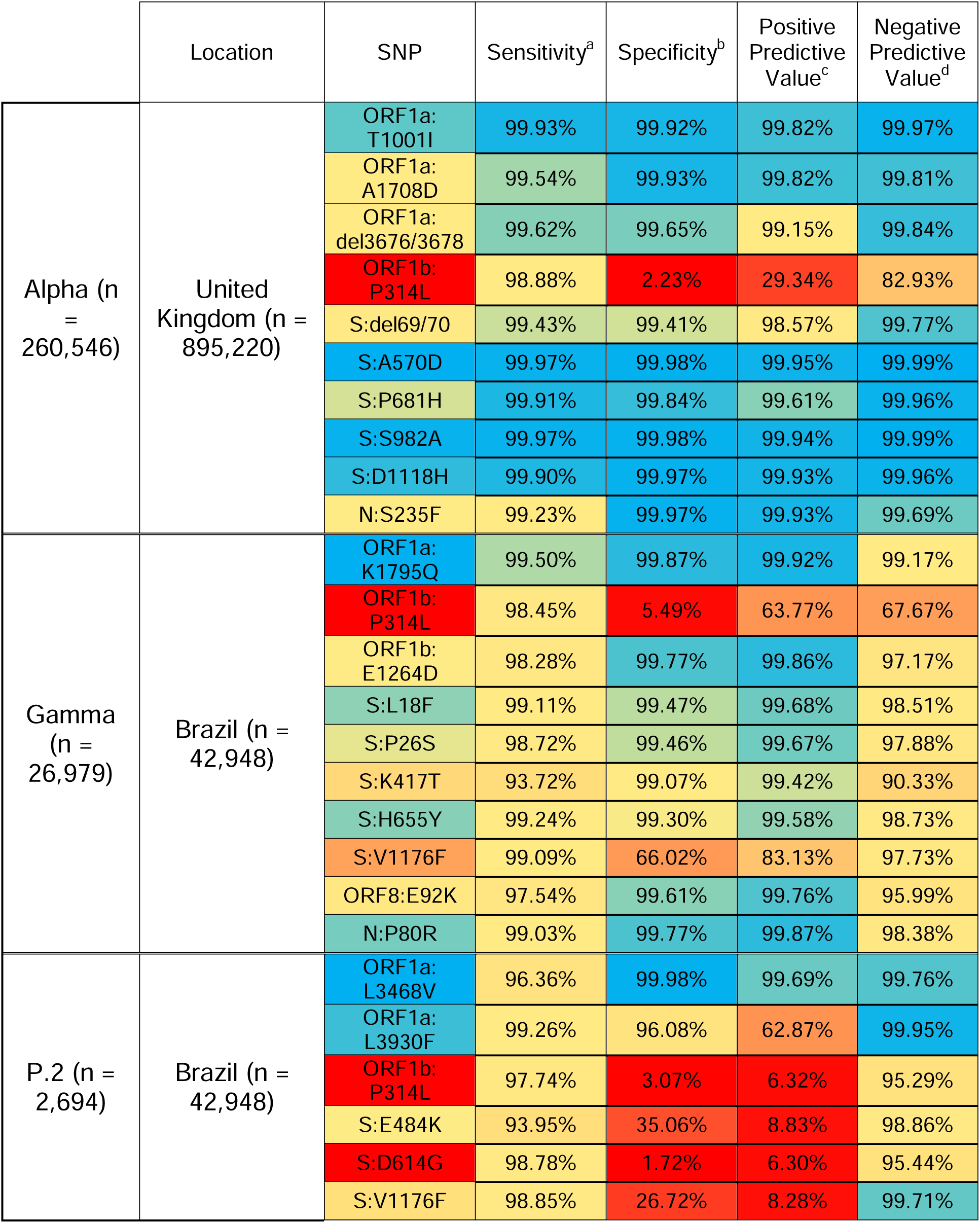
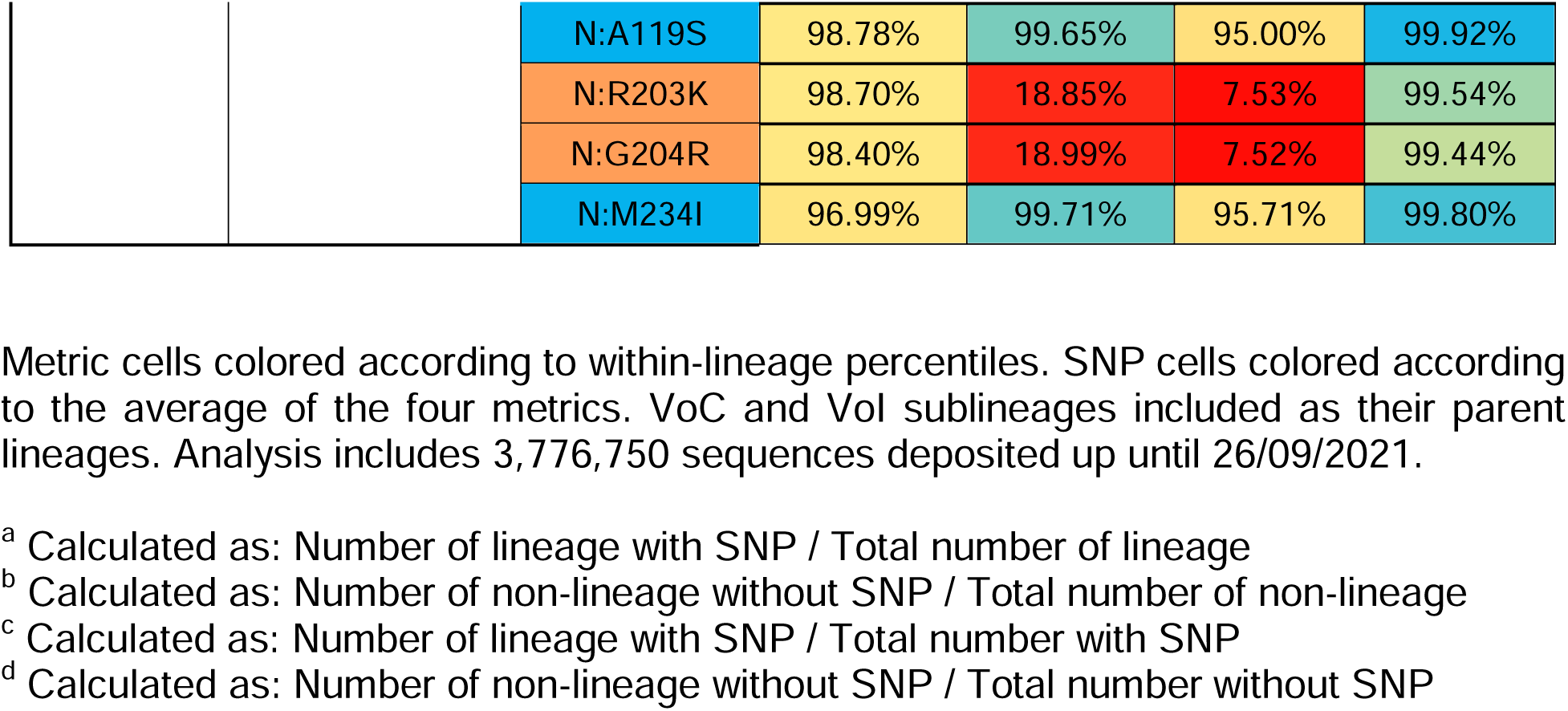
Country-Specific Performance of Lineage-Defining SNPs.

Analyses of potential target SNPS at a global scale for Alpha, Gamma, P.2, and other VoCs and VoIs are included in the supplementary material as reference (Supplementary Table 3).

### ASP-PCR and NGS performance

RNA was extracted from primary oropharyngeal or nasopharyngeal samples from participants in the REMAP-CAP and COV003 trials with confirmed SARS-CoV-2 infection and processed via either ASP-PCR, NGS, or both methods. For REMAP-CAP, this included 717 samples for ASP-PCR and 1,130 samples for NGS. For COV003, this included 365 samples for ASP-PCR and 641 samples for NGS. NGS results were assessed either for their ability to produce a lineage using the Pangolin software^15^ (NGS – Pangolin) or base calling over the SNP(s) targeted by ASP-PCR (NGS – SNP).

As the range of viral loads between the trials and methods varied, direct comparisons were achieved via deriving probit regression models for both methods and trials to predict the likelihood of producing a lineage designation. The probit results were visualized by applying the resultant functions to a dataset of 10,000 randomly generated viral loads. From this, the influence of sample viral load on method success was measured (Figures 1 & 2).

**Figure 1).**
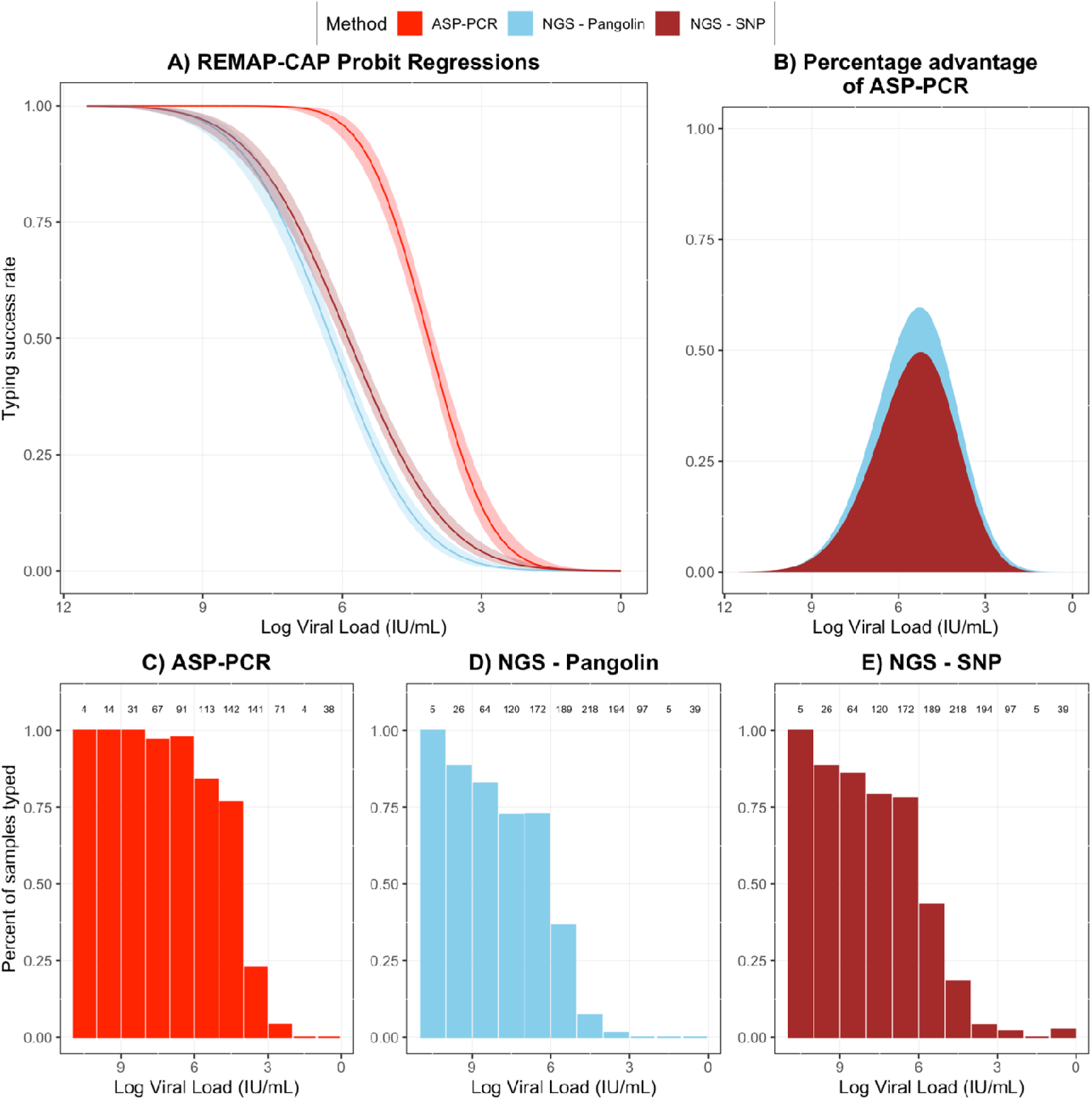
Performance of ASP-PCR and NGS in REMAP-CAP trial. Panel A) Probit regression of likelihood of lineage designation success for ASP-PCR, NGS - SNP, and NGS - Pangolin derived from REMAP-CAP samples. B) Percentage difference between probit regression typing success rates for ASP-PCR vs NGS – SNP or NGS – Pangolin. C-E) Individual method performance on REMAP-CAP samples. Fractions on top indicate total samples in one log data bins.

**Figure 2).**
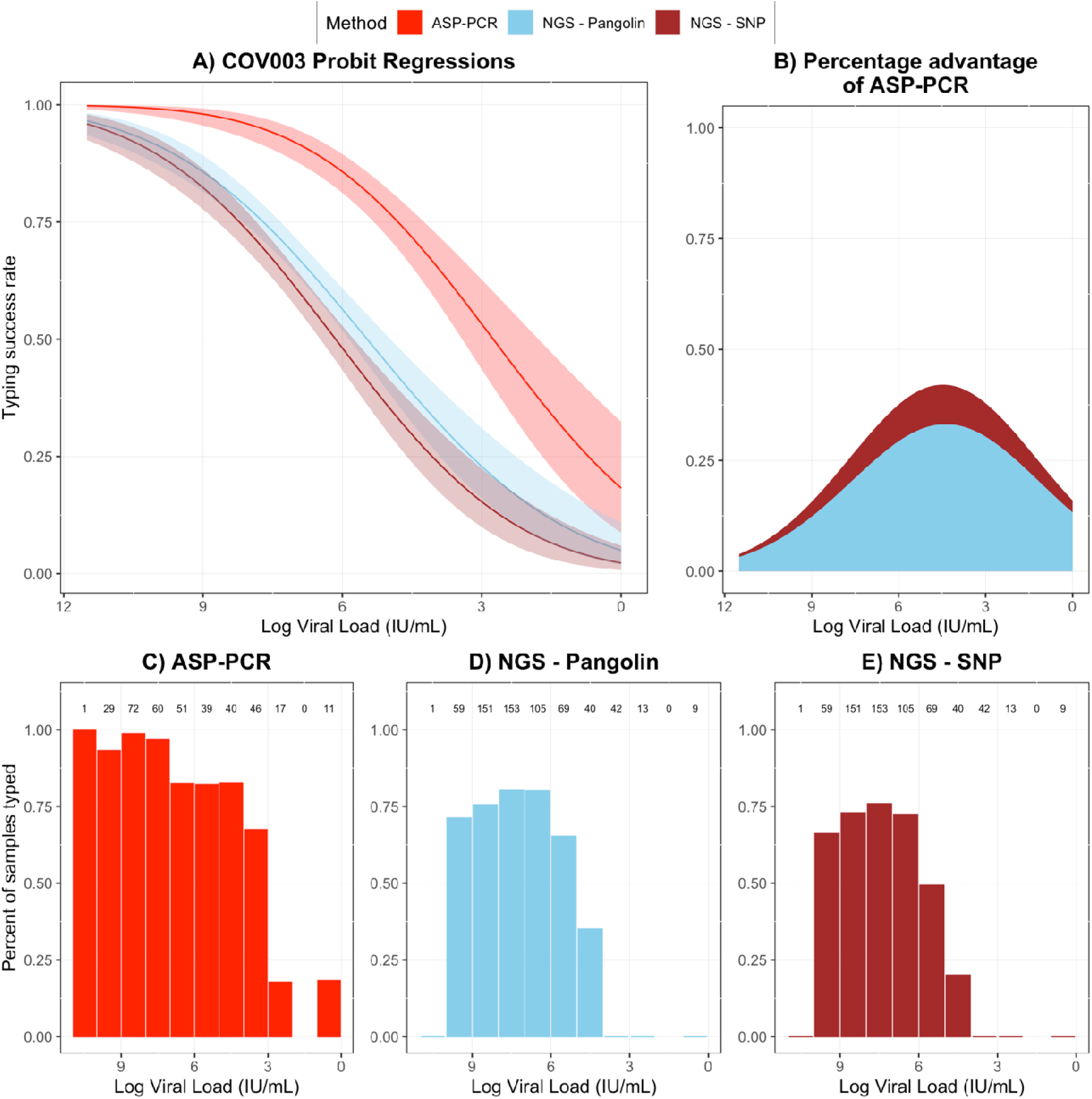
Performance of ASP-PCR and NGS in COV003 trial. Panel A) Probit regression of likelihood of lineage designation success for ASP-PCR, NGS - SNP, and NGS - Pangolin derived from COV003 samples with 95% confidence intervals. B) Percentage difference between probit regression typing success rates for ASP-PCR vs NGS – SNP or NGS – Pangolin. C-E) Individual method performance on COV003 samples. Numbers above indicate total samples in one log data bins.

In the REMAP-CAP trial, ASP-PCR had a >25% greater likelihood of producing a lineage designation over NGS – Pangolin for viral loads in the range of 2.26 * 10^3^ – 1.79 * 10^7^ IU/mL and over NGS – SNP for viral loads in the range of 5.48 * 10^3^ – 9.38 * 10^6^ IU/mL (Figure 1b). These ranges capture 61.7% and 51.4%, respectively, of the 1,289 total samples quantified in the trial. In COV003, a >25% greater likelihood of producing a lineage designation over NGS – Pangolin was seen for the viral load range of 8.53 * 10^1^ – 6.90 * 10^6^ IU/mL and over NGS – SNP for viral loads in the range of 1.56 * 10^1^ – 5.77 * 10^7^ IU/mL (Figure 1b), a wider range than for REMAP-CAP but peaking at a lower maximum percentage difference. For COV003, for which the distribution of viral loads was more skewed to higher values, these ranges capture 39.8% and 59.6%, respectively, of the viral loads of the 653 total samples quantified by RT-qPCR. At functionally all viral loads in both trials, ASP-PCR was superior to NGS for making a lineage designation with NGS – SNP/Pangolin only marginally outperforming ASP-PCR at extreme viral loads not seen in samples quantified in the trials.

Concerns regarding the quality of RNA from several recruitment sites in the COV003 trial motivated by short average read lengths prompted investigation of the utility of ASP-PCR for degraded samples. Degraded samples were defined as those that returned a viral load estimate >10^6^ IU/mL (Ct value ∼26 or less in CDC N1 RT-qPCR assay) but fewer than 20,000 bases with a read depth of at least two reads. Subsetting analysis to these samples (n = 113 for ASP-PCR, n = 118 for NGS – Pangolin and NGS – SNP) or those that were nondegraded (all samples that did not meet criteria for degradation; n = 252 for ASP-PCR, n = 523 for NGS – Pangolin and NGS – SNP), the advantages of ASP-PCR are clear (Figure 3). ASP-PCR was minimally impacted by the sample degradation status successfully typing 95.6% of the 113 total degraded samples tested by ASP-PCR, while the utility of both NGS – Pangolin and NGS – SNP were massively reduced, only typing 11.0% and 0.0% of the 118 degraded samples processed by NGS, respectively.

**Figure 3).**
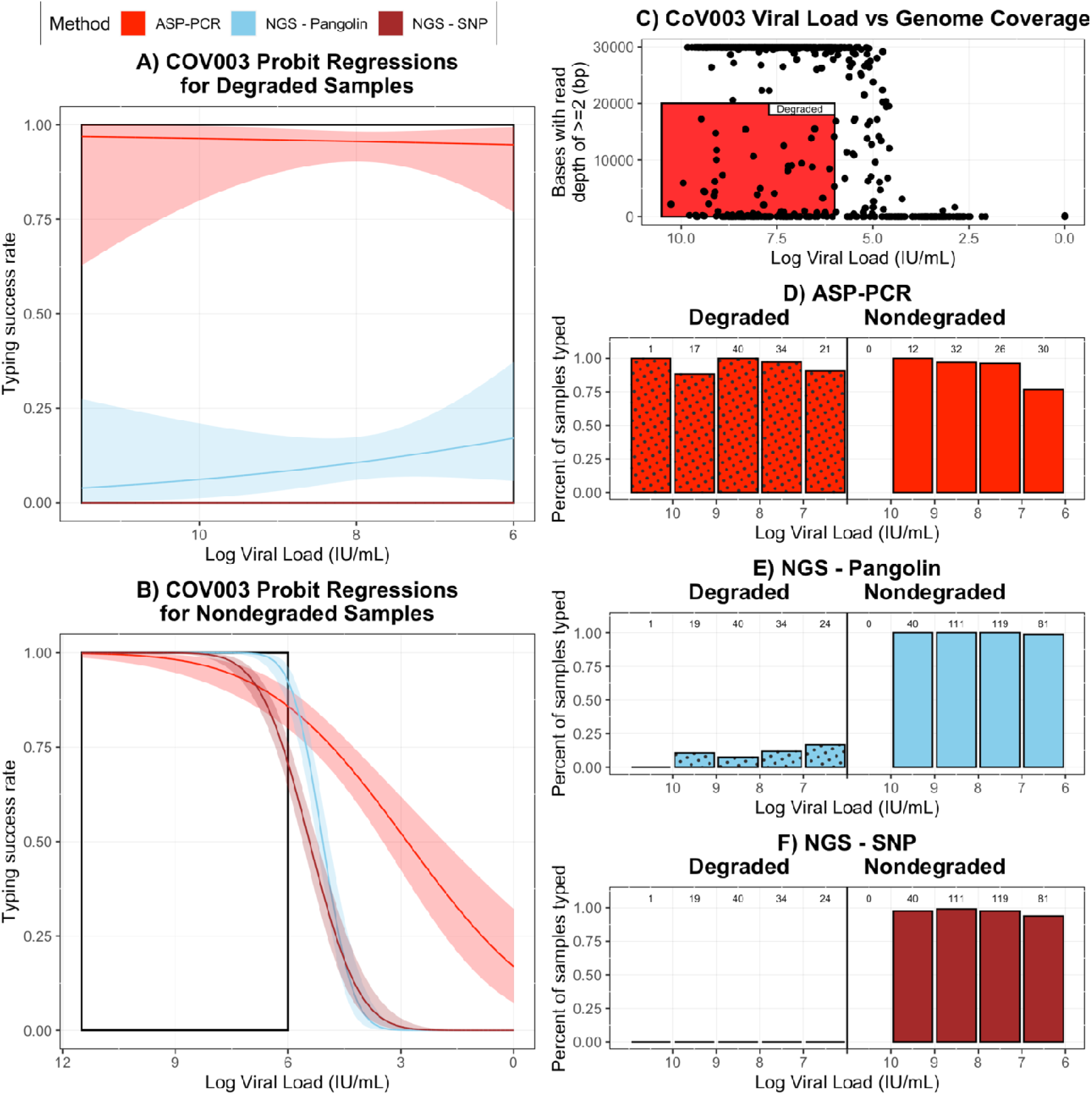
Impact of Degraded RNA in COV003 Samples on Method Performance. Panel A) Probit regression of likelihood of lineage designation success for ASP-PCR, NGS - SNP, and NGS - Pangolin derived from degraded COV003 samples with 95% confidence intervals. NGS - SNP failed to type any samples with low genome coverage. B) Probit regression of likelihood of lineage designation success for ASP-PCR, NGS - SNP, and NGS - Pangolin derived from nondegraded COV003 samples with 95% confidence intervals. For A) and B), black box indicates viral loads >10^6^ IU/mL. C) Genome coverage of COV003 samples plotted vs sample viral load. Samples with <= 20,000 bases with >= 2 reads and viral load > 10^6^ IU/mL were defined as degraded (red box). D-F) Individual method performance on COV003 degraded and nondegraded samples. Numbers above indicate total samples in one log data bins.

For the 181 REMAP-CAP samples with paired ASP-PCR and NGS - Pangolin results, lineage designation was concordant for 99% (180/181) samples. A single sample was called Alpha by ASP-PCR but the parent lineage B.1.1 by NGS - Pangolin – the sequence data indicated S:1118D. For the 122 COV003 samples with paired ASP-PCR and NGS - Pangolin results, designations were concordant for 85% (104/122) of samples. All 18 discordant samples were called Gamma or P.2 by the ASP-PCR. However, this discrepancy was likely due to miscalls by Pangolin, as 17/18 of these discordant samples were typed as Gamma or P.2 by phylogenetic reconstruction (supplementary methods), leading to a more accurate assessment of ASP-PCR and NGS concordance for COV003 of 99% (121/122).

Validation data for ASP-PCR has been presented elsewhere.^10,16^ Briefly, against a panel of 40 known samples of varying viral loads, S:D118H/Alpha demonstrated 97.5% sensitivity and 100% specificity, S:K417T/Gamma demonstrated 90% sensitivity and 95% specificity for 49 samples, and ORF1a:L3468V demonstrated 100% sensitivity and 91% specificity for 47 samples.

### Predicted performance

The output of a probit regression is a link function describing the likelihood of a binary outcome for a given set of input variables. As such, the function can be applied over a dataset containing the input variable of interest to predict the success over a sample population. To estimate the utility of ASP-PCR over other study populations, the REMAP-CAP S:D1118H/Alpha ASP-PCR, NGS - Pangolin, and NGS - SNP probit regression models were applied over published datasets containing Ct or viral load data (Table 3).^17,18^ The advantage of ASP-PCR seen in both the hospitalized and community testing cohorts was driven by large proportions of samples in the Ct 26-30 range. Focusing on the subset of samples that fall in that range in the Tso et al. (2021) dataset, ASP-PCR was predicted to successfully type 95.7% of the samples versus only 40.9% by NGS – SNP and 30.5% by NGS – Pangolin.

### Effect of molecular modifications

To measure the effects of the addition of low molecular amides, oligonucleotides modified with LNAs, or both treatments, a panel of 83 samples of varying viral load from REMAP-CAP were re-extracted from the primary samples and used to screen effects on ASP-PCR performance. In samples untreated with 2-pyrrolidinone, modification of oligonucleotides to LNAs was associated with a statistically significant increase in typing percentage (p = 0.04, Fisher’s Exact Test; Figure 4). The general trend was to increased typing percentage with LNAs and decreased with 2-pyrrolidinone, but the other comparisons did not reach significance.

**Figure 4).**
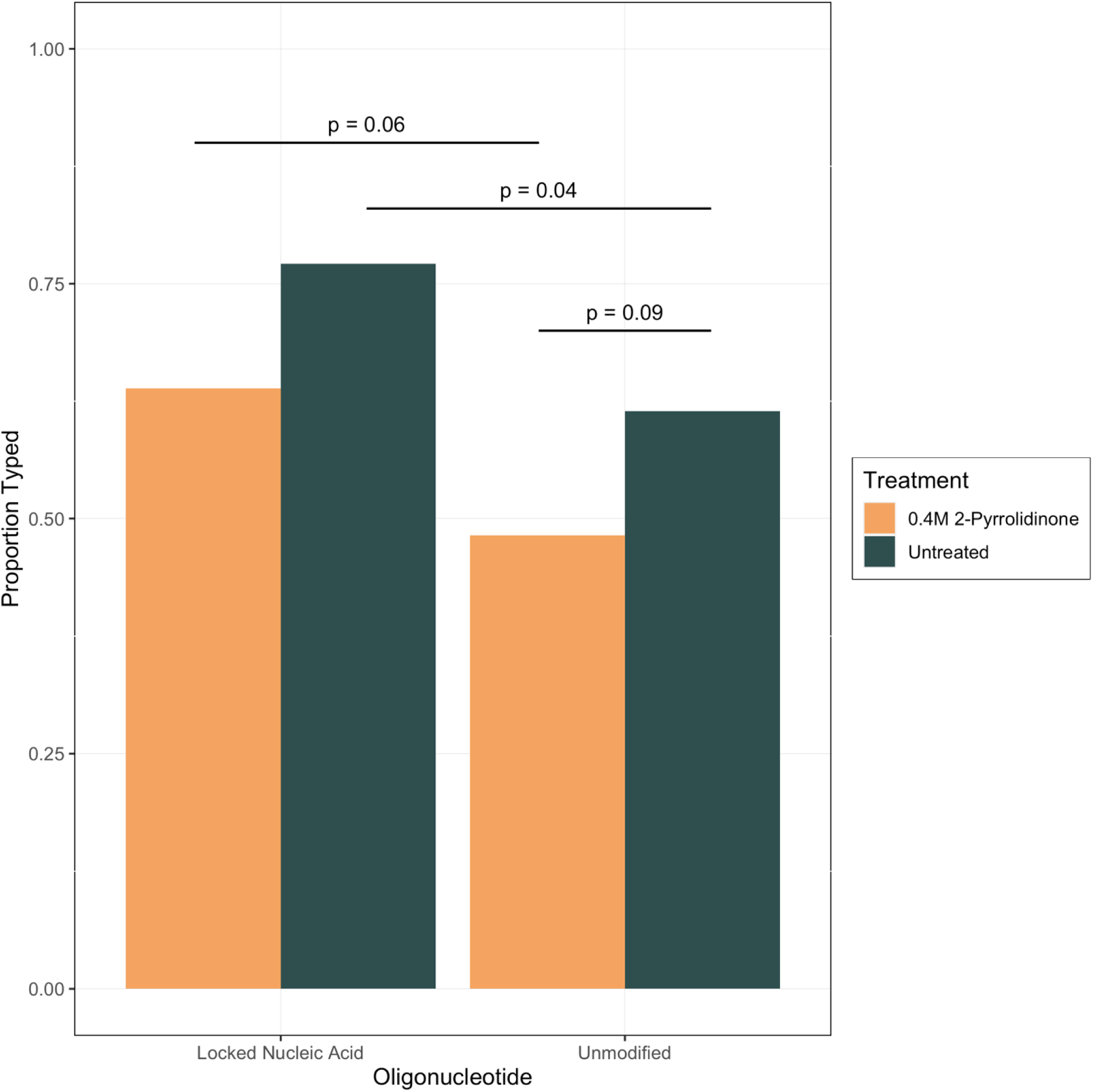
Effect of Molecular Modification on S:D1118H/Alpha ASP-PCR performance. Percentage of total samples successfully typed using approach described in materials and methods. P values from Fisher’s Exact Test. Lineage designation (e.g., WT or Alpha/B.1.1.7) were concordant for all samples for all methods.

While the addition of 0.4M 2-pyrrolidinone did decrease the raw signal observed for the discriminating probe, this improvement in discrimination came at the expense of lineage designation for viral load samples of all concentrations (Supplementary Figures 2, 3). Additionally, LNA probes returned signals for samples negative by N gene qPCR, which may represent false positives, although the confidence intervals on the probit regressions are wide.

## DISCUSSION

The benefits of NGS extend far beyond lineage designation and can be applied to answer numerous research questions of public health importance including identifying genomic sites under positive selection, in-depth molecular epidemiology, and detection of novel variants.^1^ Perhaps most importantly to this project, the design of ASP-PCR oligonucleotides and the very knowledge of what lineages to target is entirely dependent on the generation and dissemination of SARS-CoV-2 sequences produced via NGS. However, as evidenced here, there exist multiple use cases where ASP-PCR could be applied to improve variant identification. In general, ASP-PCR would be best deployed in studies wherein the lineages of interest are known; the expected frequency of the target lineage in the study population can be estimated; and there are not co-circulating SARS-CoV-2 variants that contain the target SNP for the study period.

Appropriate SNP selection is the key to the interpretability of ASP-PCR data. The global PPV and NPV estimates presented in supplementary table 3 are biased due to the analysis: a) assuming random selection of any sequence that has been sequenced during the pandemic (ignoring geographic and temporal influences); and b) not factoring in biases towards sequencing coverage per country being nonrandom (e.g., the United Kingdom provided 23.7% of sequencing data in the analysis). Geographic restrictions for sequence analysis improve estimates of PPV and NPV and could allow use of targets that appear less suitable in the global analysis. For example, if one restricts analysis of the S:K417N mutation for Beta to only South African sequences, the PPV increases from 82.23% to 95.27% (Supplementary Table 3). This impact is also demonstrated in directly comparing table 1 and supplementary table 3. In contrast, the global sensitivity and specificity values are less susceptible to these assumptions and should be the starting point for designing novel ASP-PCR oligonucleotide sets before refinement using temporally and location-specific restrictions.

Practically, ASP-PCR may also prove a useful alternative for analyzing stored samples where volumes or viral loads are too low to be used as inputs into NGS platforms. As shown in figure 1A and table 3, ASP-PCR may be most appropriately used in settings where patients may present with lower viral loads and SARS-CoV-2 lineage data may provide useful post-hoc analysis of therapeutic efficacy. Further, as demonstrated in our previous work,^10^ the rate of appearance of a specific lineage in hospitalized patients could be compared to that in the general population to estimate increased severity. The method also presents as particularly useful for analyzing samples suffering from known or suspected degradation for which any NGS method would likely be inviable. Lastly, ASP-PCR may be an attractive alternative to NGS in resource-limited areas due to its reduced reagent costs, use of equipment available in most microbiology labs, lower standard of technical expertise needed, and quicker turnaround in generating data.

Other attempts have been made to leverage RT-PCR to identify SARS-CoV-2 variants. Lee et al. (2021) designed an Alpha/B.1.1.7 allele-specific PCR based on the S:H69/V70del, S:Y144del, and S:A570D mutations with mismatches in the 3’ end of the forward primer.^19^ A weakness of their approach is that at least two independent AS-PCR reactions are needed to perform lineage identification for individual samples, although the method could be used to estimate lineage prevalence for pooled samples as described in the study. Babiker, et al. (2021) constructed a multiplex, single reaction ASP-PCR based on SNP identification from dropout of signal for probes targeting S:K417 and the S:E484K and S:N501Y mutations, although these are likely uninformative for variant identification (see Supplementary Table 3).^20^ The Vogels et al. (2021) multiplex RT-qPCR method discriminates Alpha, Beta, Gamma, and other by targeting the deletions ORF1a:3675-3677del and S:69/70del.^21^ Lastly, Harper et al. (2021) designed a genotyping panel for identifying 19 SNPS based off of PACE chemistry and allele-specific forward primers.^22^ Lee et al. (2021) and Harper et al. (2021) both based their designs on 3’ mismatches being lethal for PCR reactions, but experimental evidence challenges that assumption.^23^ In addition, several commercial companies are now offering SARS-CoV-2 PCR assays based on the principles of ASP-PCR, but researchers will typically require knowledge of which SNPs have been targeted and their specificity for strain identification in their research areas. They may find greater cost-effectiveness and adaptability with in-house designed oligonucleotides.

The drop in performance for some high viral load samples for both NGS methods in COV003 versus REMAP-CAP was thought to be due to fragmentation of RNA during sample storage, as evidenced by shorter than expected library insert size, high duplication rates, and variable performance on samples from different recruitment sites.^16^ The RT-qPCR and ASP-PCR amplicon length likely impacted their respective performance for COV003 samples, as highly fragmented RNA may not have had intact RNA spanning the primer binding sites. For COV003, the shorter amplicon (S:K417T/Gamma) performed slightly better for high viral load samples, but the confidence intervals overlapped for all viral loads (Supplementary Figure 1). The ability of ASP-PCR to, on rare occasions, designate lineages on samples negative via N gene qPCR in the COV003 trial but not the REMAP-CAP trial is likely also due to this.

One major benefit of ASP-PCR is the flexibility and adaptability to target new SARS-CoV-2 SNPs as they arise. In our experience, SNP targets should fulfill most or all of the following criteria to ease design and optimization:

- Follow best practices for primer/probe design and amplicon length as specified by the RT-PCR mix being used. Place the target base in or near the middle of the probe.
- If possible, choose a target SNP with an adenine/uracil up- and downstream of the mismatched base to increase the energetic cost of the mismatched base.
- In contrast to RT-qPCR where the T_M_ of the probe should be several degrees higher than primers, all oligonucleotides for ASP-PCR should be designed to melt at similar temperatures.
- Optimization should begin with the annealing/extending temperature ∼5°C below the lowest T_M_ of the two probes. Decrease the temperature to increase raw signal; increase the temperature to increase discrimination. Reaction conditions cannot easily be predicted computationally and can most readily be experimentally confirmed using cDNA aliquots of sequence confirmed samples.
- Label both probes with bright fluorophores, such as 6-FAM and HEX.

Previous research on improving the specificity of PCR has shown that addition of low-molecular weight amides can reduce the amplification of off-target PCR products.^24^ It is theorized that the mechanism of action for organic additives is destabilization of template double-helices;^25^ this effect would partially explain the drops in raw signal seen in samples treated with 2-pyrrolidinone and decreased amplification for most samples. Moving forward, modifications that increase the relative destabilization caused by a single base mismatch (as with locked nucleic acids) rather than disrupting binding dynamics of all oligonucleotides (as with 2-pyrrolidinone) should be prioritized.

ASP-PCR is a highly accurate and sensitive method for discriminating SARS-CoV-2 lineages and its application to support specific research aims is well supported by the data presented in this study.

## MATERIALS AND METHODS

### Study Design

Samples used for method development and assessment in this study were collected as part of two clinical trials conducted during periods of VoC and VoI emergence in their study locations. The variants were hypothesized to have an impact on outcomes seen within these trial; thus, highly sensitive methods for identifying specific variants were sought and developed. All samples were processed by ASP-PCR, NGS, or both methods and no sample results were excluded from this study.

### Sample Collection

The REMAP-CAP Immunological Domain was an international, open-label, randomized convalescent plasma trial enrolling patients aged 18 years or older receiving intensive care-level organ support. Patients were eligible given admission to intensive care within 48 hours of hospital admission and a positive SARS-CoV-2 microbiological test. A full trial protocol and efficacy results are available elsewhere.^7,8^ Oropharyngeal or nasopharyngeal swabs were taken prior to randomization, transported to a central academic hospital and frozen at -80°C, shipped on dry ice to a central testing laboratory, and processed as described below.

COV003 is a participant-blinded, randomized, controlled phase 3 multi-site trial that began in Brazil on June 23, 2020 assessing the efficacy of the ChAdOx1 nCoV-19 vaccine against symptomatic SARS-CoV-2 infection. Efficacy, safety data, the full study protocol, and exploratory analysis of lineage-specific efficacy are available elsewhere.^16,26,27^ Volunteers recruited for the trial were 18 years or older and at high risk of exposure (e.g., healthcare workers). Volunteers who developed primary COVID-19 symptoms were asked to contact their study site. Nasopharyngeal swab samples that were separately confirmed SARS-CoV-2 positive using commercial NAAT assays at local laboratories were shipped to a central testing laboratory and processed as described below.

### Ethics statement

REMAP-CAP and COV003 were conducted according to the principles of the latest version of the Declaration of Helsinki (version Fortaleza 2013). REMAP-CAP was performed in accordance with regulatory and legal requirements (EudraCT number: 2015-002340-14) and was approved by London-Surrey Borders Research Ethics Committee London Centre (18/LO/0660). COV003 was approved by the Brazilian National Research Ethics Committee (ref: 32604920.5.0000.5505) and the Oxford Tropical Research Ethics Committee (ref: 20-36).

### Nucleic Acid Extraction

REMAP-CAP samples and research reagent 19/304 (National Institute of Biological Standards and Control (NIBSC)) containing encapsulated, quantified full-length SARS-CoV-2 RNA were extracted using either the QIAamp Viral RNA Mini Kit (QIAGEN) as described previously or the Quick-DNA/RNA Viral Kit (Zymo Research).^10^ COV003 samples were extracted using the Quick-DNA/RNA Viral Kit as described previously.^16^

### Real-Time Quantitative PCR

SARS-CoV-2 viral RNA was detected and quantified by real-time quantitative PCR (RT-qPCR) as previously described using oligonucleotides listed in Supplementary Table 1 (ATDBio).^10^ SARS-CoV-2 RNA was quantified using a standard curve of research reagent 19/304 serially diluted from 10,000 copies/reaction to 100 copies/reaction (REMAP-CAP) or 1,000 copies/reaction to 10 copies/reaction (COV003). RT-qPCR cycle threshold (Ct) values were converted to copy number/reaction by use of the standard curve and to international units (IU)/mL by the conversion rate in the product sheet.

### ASP-PCR

SNP sites targeted for lineage discrimination were chosen based on their lineage-specific predictive value estimated using publicly available sequence data published on GISAID.^13^

ASP-PCR was performed using the Quantitect Probe RT-PCR kit (QIAGEN) with 5 µL of extracted RNA in a 25 µL reaction volume on an Applied Biosystem StepOnePlus Real-Time PCR system using the genotyping program. SNPs were designated based on their clustering with discrimination controls. Serially diluted cDNA aliquots of sequence-confirmed samples were used as discrimination controls; ultrapure water served as negative controls. Samples that failed to achieve a change in signal in either probe greater than those of the no template controls or lacked evidence of amplification were designated “undetermined”. Reaction conditions (annealing and extension temperature and length and oligonucleotide concentrations) were optimized using serially diluted cDNA generated from samples of known lineage. The RT-PCR settings for each ASP-PCR are described in supplementary table 2.

**Table 2).** Predicted Performance of S:D1118H/Alpha ASP-PCR, NGS - SNP, and NGS - Pangolin.

| Study description | Samples used in analysis | n | Viral load variable | Mean (SD) | Predicted ASP-PCR performance (95% CI) | Predicted NGS – SNP performance (95% CI) | Predicted NGS – Pangolin performance (95% CI) |
| --- | --- | --- | --- | --- | --- | --- | --- |
| Patients hospitalized with COVID-19 sampled within 48 hours of hospital admission in New York City, NY<sup>17</sup> | All samples | 1,044 | Ct, unclear gene target | 27.63 (6.18) | 74.1% (72.1%, 76.0%) | 47.2% (44.2%, 50.1%) | 41.8% (39.0%, 44.5%) |
| Community testing in Texas<sup>18</sup> | All samples | 57,517 | N gene Ct | 22.90 (5.97) | 96.3% (94.9%, 97.3%) | 67.9% (65.0%, 70.6%) | 62.9% (60.0%, 65.6%) |
|  | Samples Ct < 26 | 34,718 |  | 18.83 (4.38) | 99.9% (99.9%, 100.0%) | 88.6% (86.0%, 90.7%) | 86.9% (84.3%, 89.1%) |
|  | Samples Ct >= 26 & < 30 | 17,567 |  | 28.20 (1.10) | 95.7% (92.9%, 97.5%) | 40.9% (37.4%, 44.5%) | 30.5% (27.1%, 34.1%) |
|  | Samples Ct > 30 | 5,232 |  | 31.05 (0.82) | 73.4% (67.9%, 78.4%) | 21.7% (18.7%, 24.9%) | 12.3% (9.87%, 15.2%) |

To test the effects of modified oligonucleotides, the D1118H/Alpha ASP oligonucleotide set was redesigned using locked nucleic acids (LNA) over the SNP site (Supplementary Table 1). To measure the impact of low-molecular weight amides on differentiation, 2-pyrrolidinone (Merck) was added to the qPCR master mix at a reaction concentration of 0.4M as recommended in Chakrabarti and Schutt (2001).^24^

### Next-Generation Sequencing

Samples were sequenced using the veSEQ NGS protocol as described previously.^2^ An extended NGS method description is available in the supplementary material. Two approaches were used to assess lineage assignment by veSeq. NGS - SNP evaluated variant calls at the same sites of interest targeted by ASP-PCR. Consensus sequences with no coverage over the site of interest were deemed “undetermined”. NGS - Pangolin assessed the ability of the widely used Pangolin tool (v3.1.11 for REMAP-CAP and v.2.4.2 for COV003) to make a lineage designation (specifically, testing whether sequences meet the default quality control).^15^ Degraded samples were defined as those with high viral loads (>=10^6^ IU/mL) but poor genome coverages (<=20,000 bases with read depth of at least two reads).

### Data analysis

Probit regression models were generated using base function ‘glm’ in R version 4.0.4.^28^ Model outputs are available in the supplementary material. Confidence intervals were generated in consultation with open-source code and methods published by Dr. Gavin Simpson on his website.^29^

### Predictive Performance

Method performance on representative datasets was estimated by summing the probit function output for each individual sample. IU/mL, N gene Ct value, or average Ct value were used as the model input variable depending on metadata availability.

### Data availability

Data used to generate figures and statistical analysis in this manuscript are available from the corresponding author at reasonable request.

## Data Availability

All data produced in the present study are available upon reasonable request to the authors

## Acknowledgements

The views expressed in this publication are those of the authors and not necessarily those of any funding bodies. We thank all volunteers who supported both studies presented in this article. JR is supported by Marshall and Clarendon Scholarships. MS-H is supported by the National Institute for Health Research Clinician Scientist Award (NIHR-CS-2016-16-011). We thank Professor James Szymanski and Dr. Chak Foon Tso and the team at Dascena for the generous provision of raw viral load data from their respective studies.

## Funding

The COV003 trial is supported by the National Institute of Health Research (NIHR), the Lemann Foundation, Rede D’Or, the Brava and Telles Foundation, and AstraZeneca. The REMAP-CAP trial is supported by The Platform for European Preparedness Against (Re-) emerging Epidemics (PREPARE) consortium by the European Union, FP7-HEALTH-2013-INNOVATION-1 (#602525), the Australian National Health and Medical Research Council (#APP1101719), the Australian Medical Research Future Fund (#APP2002132), the New Zealand Health Research Council (#16/631), the Canadian Institutes of Health Research COVID-19 Rapid Research Funding Grant (#447335), the Canadian Institute of Health Research Strategy for Patient-Oriented Research Innovative Clinical Trials Program Grant (#158584), the National Institute for Health Research (UKRIDHSC COVID-19 Rapid Response Rolling Call, “The use of convalescent plasma to treat hospitalised and critically ill patients with COVID-19 disease” (COV19-RECPLAS), the UK National Institute for Health Research (NIHR) and the NIHR Imperial Biomedical Research Centre, the Health Research Board of Ireland (CTN 2014-012), the UPMC Learning While Doing Program, the Translational Breast Cancer Research Consortium, the Pittsburgh Foundation, the French Ministry of Health (PHRC-20-0147), the Minderoo Foundation, and the Wellcome Trust Innovations Project (215522). Australian governments fund Australian Red Cross Lifeblood for the provision of blood products and services to Australia. Collection of UK plasma was funded by the DHSC through core funding under COVID-19 and EU SoHo Grants.

## Author Contributions

Conceptualization: JR, PS

Methodology: DB, TG, HH, JR, PS

Software: TG, JR

Formal Analysis: TG, JR, PS

Investigation: CJ, GMC, DN, JR, SW

Resources: FA, SB, SACC, LE, AE, MF, PMF, ACG, AJ, EL, TL, DM, PRM, AJP, MNR, DJR, KMR, JR, MSH

Data Curation: TG, JR, PS

Writing – Original Draft: JR

Writing – Review & Editing: SB, HH, TG, CJ, JR, AJP, MNR, MSH, PS

Supervision: TG, HH, PS

## Declaration of Interests

The funders of REMAP-CAP had no role in the design, analysis, or interpretation of the data presented in this study. Oxford University has entered into a partnership with AstraZeneca for further development of ChAdOx1 nCoV-19. AstraZeneca did not have a role in reviewing the data for this study. AJP is an NIHR senior investigator.

## Notes

### Competing Interest Statement

Oxford University has entered into a partnership with AstraZeneca for further development of ChAd-Ox1 nCoV-19. AstraZeneca did not have a role in reviewing the data for this study.

### Clinical Trial

NCT02735707 (REMAP-CAP)
NCT04536051 (COV003)

### Author Declarations

REMAP-CAP was performed in accordance with regulatory and legal requirements (EudraCT number: 2015-002340-14) and was approved by London-Surrey Borders Research Ethics Committee London Centre (18/LO/0660). COV003 was approved by the Brazilian National Research Ethics Committee (ref: 32604920.5.0000.5505) and the Oxford Tropical Research Ethics Committee (ref: 20-36).

